# Elevated Maternal Glycated Albumin is Associated with Large for Gestational Age Infants: Results from a Prospective Longitudinal Study

**DOI:** 10.64898/2026.11.02.26367057

**Authors:** Carolina Ferreira Jørgensen, Thilo Samson Chillon, Cathrine Scavenius, David B. Sacks, Bharath Raghuraman, Dorte Bekker-Nielsen, Nicolai Bache, Ole Vorm, Line Rode, Charlotte Ekelund, Peter Damm, Kurt Højlund, Dorte Møller Jensen, Tine Dalsgaard Clausen, H. David McIntyre, Martin Overgaard

## Abstract

**Aims/hypothesis:** Glycated albumin (GA) has been shown to be a moderate predictor of gestational diabetes mellitus (GDM), while evidence supporting its role in predicting adverse pregnancy outcomes remains limited. Here, we aimed to perform a comparative study of GA and haemoglobin A1c (HbA_1c_) for predicting GDM and large-for-gestational-age (LGA) infants in a prospective cohort.

**Methods:** We developed a rapid liquid chromatography-tandem mass spectrometry method for measuring site-specific albumin glycation. GA was quantified at two glycation sites, GA (KQT) and GA (KLV), as well as their sum, GA-total. GA was measured in stored serum samples collected at 11-14 and 23-31 gestational weeks from 465 participants in the single-centre observational cohort. HbA_1c_ was measured routinely at each visit. Linear mixed-effects models were used to compare GA between participants with and without GDM (n=100) or LGA (n=57). Logistic regression was used to estimate odds ratios (ORs) for GDM and LGA according to GA and HbA_1c_.

**Results:** GA was lower in women who developed GDM during early pregnancy compared to non-GDM controls (GA-total; *p*=0.004), while in mid-late pregnancy, only GA (KQT) was lower in women with GDM (*p*=0.036). In contrast, HbA_1c_ was higher in women with GDM at both timepoints compared to non-GDM controls (*p*<0.001) and higher HbA_1c_ was associated with increased odds of GDM (OR=2.20, *p*<0.001; OR=2.30, *p*<0.001, respectively). GA was higher in the LGA group, in both early (GA-total; *p*=0.032) and mid-late pregnancy (GA-total; *p*=0.030) and associated with increased odds of LGA (GA-total; OR=1.59, *p*=0.009; GA-total; OR=1.46, *p*=0.019, respectively). HbA_1c_ was not associated with LGA at either timepoint.

**Conclusions/interpretation:** Higher GA in early and mid-late pregnancy were associated with an increased risk of LGA, whereas no significant association was observed for HbA_1c_. GA may therefore provide risk assessment beyond HbA_1c_ for identifying pregnancies at risk of fetal overgrowth.

**Research in Context:** *What is already known about this subject?:*

- Glycated albumin (GA) has been proposed as a biomarker for GDM, LGA, and other adverse pregnancy outcomes.
- HbA_1c_ is an established marker of chronic glycaemia outside pregnancy, but its role in the routine diagnosis and monitoring of GDM during pregnancy remains limited.
- Both GA and HbA_1c_ are markers of glycaemic exposure; however, they capture different time frames and are influenced differently by the physiological changes of pregnancy.

*What is the key question?:*

- Are GA and HbA_1c_ measured during pregnancy associated with GDM and LGA?

*What are the new findings?:*

- HbA_1c_ and GA showed opposing associations with GDM development.
- Higher GA in early and mid-late pregnancy were associated with an increased risk of LGA.
- HbA_1c_ showed limited association with LGA.

*How might this impact clinical practice in the foreseeable future?:*

- GA and HbA_1c_ may serve as complementary biomarkers for assessing glycaemia and predicting pregnancy complications, with potential utility for improving risk stratification during pregnancy.

## Introduction

Gestational diabetes mellitus (GDM) defined as hyperglycaemia with onset or first recognition during pregnancy, is one of the most common medical complications of pregnancy with a prevalence ranging from 1% to >30% [1]. GDM increases the risk of short- and long-term complications for both mother and offspring, including higher risks of caesarean section, large-for-gestational-age (LGA) infants, shoulder dystocia, neonatal hypoglycaemia, and future development of type 2 diabetes in both mother and child [2–4].

While the recommended diagnostic test for GDM includes fasting and post-load plasma glucose measured during an oral glucose tolerance test (OGTT), the long-term glycaemic marker haemoglobin A1c (HbA_1c_) has limited utility for diagnosing GDM because physiological changes during pregnancy, including increased red blood cell turnover and haemodilution, reduce HbA_1c_ independent of glycaemia [5]. In addition, HbA_1c_ primarily reflects average glycaemia over the preceding 2-3 months and is relatively insensitive to the postprandial hyperglycaemia that characterizes GDM. Consequently, HbA_1c_ lacks sufficient sensitivity for GDM diagnosis [6, 7] and is not recommended as a standalone screening or diagnostic test, although it is used early in pregnancy to identify women with previously undiagnosed overt diabetes [8, 9].

Glycated albumin (GA) reflects average glycaemia over a shorter timespan (2-3 weeks) than HbA_1c_ and has generated considerable attention as a potential early marker of GDM and adverse pregnancy outcomes [9]. While several studies have evaluated GA and HbA_1c_ for diagnosing GDM [10–12], there is a lack of evidence from prospective cohort studies exploring the associations of GA with adverse pregnancy outcomes.

Site-specific GA assays based on liquid-chromatography tandem mass spectrometry (LC-MS/MS) may provide additional molecular information by quantifying glycation at individual lysine residues, potentially revealing glycation patterns that are obscured by conventional assays which measure total GA. At present, clinical evidence supporting this approach remains limited [13].

To this end, we devised a novel high-throughput LC-MS/MS method for site-specific quantification of albumin glycation and performed a comparative study of GA and HbA_1c_ for predicting GDM and LGA in a longitudinal prospective cohort.

## Methods

### Study Design and Participation

This was a single centre observational cohort study from Hilleroed, Denmark introducing universal screening for GDM at 24-28 weeks’ gestation [14]. The study was approved by the local Committee on Health Research Ethics (H-19001203) and Data Protection Agency (P-2019-89) and was performed in alignment with the Helsinki Declaration. Women attending the first-trimester routine screening between October 2019 and March 2020 were considered for enrolment in the study. Women were deemed eligible for study participation based on eligibility and exclusion criteria as previously reported [14]. Figure 1A is an outline of the study design and population. Initially, 871 women gave full consent to participate in the study and provided a blood sample at 11-14 gestational weeks (GW) (T1). However, mainly due to COVID-19, only 465 of these women completed the OGTT and provided blood samples for the biobank at 23-31 GW (T2). Participants were analysed in two predefined comparisons: GDM vs non-GDM controls and LGA vs non-LGA controls. Among the 465 participants, 100 developed GDM and 365 were normoglycaemic controls, while 57 gave birth to an LGA neonate and 408 were non-LGA controls (Fig. 1A).

**Fig. 1.**
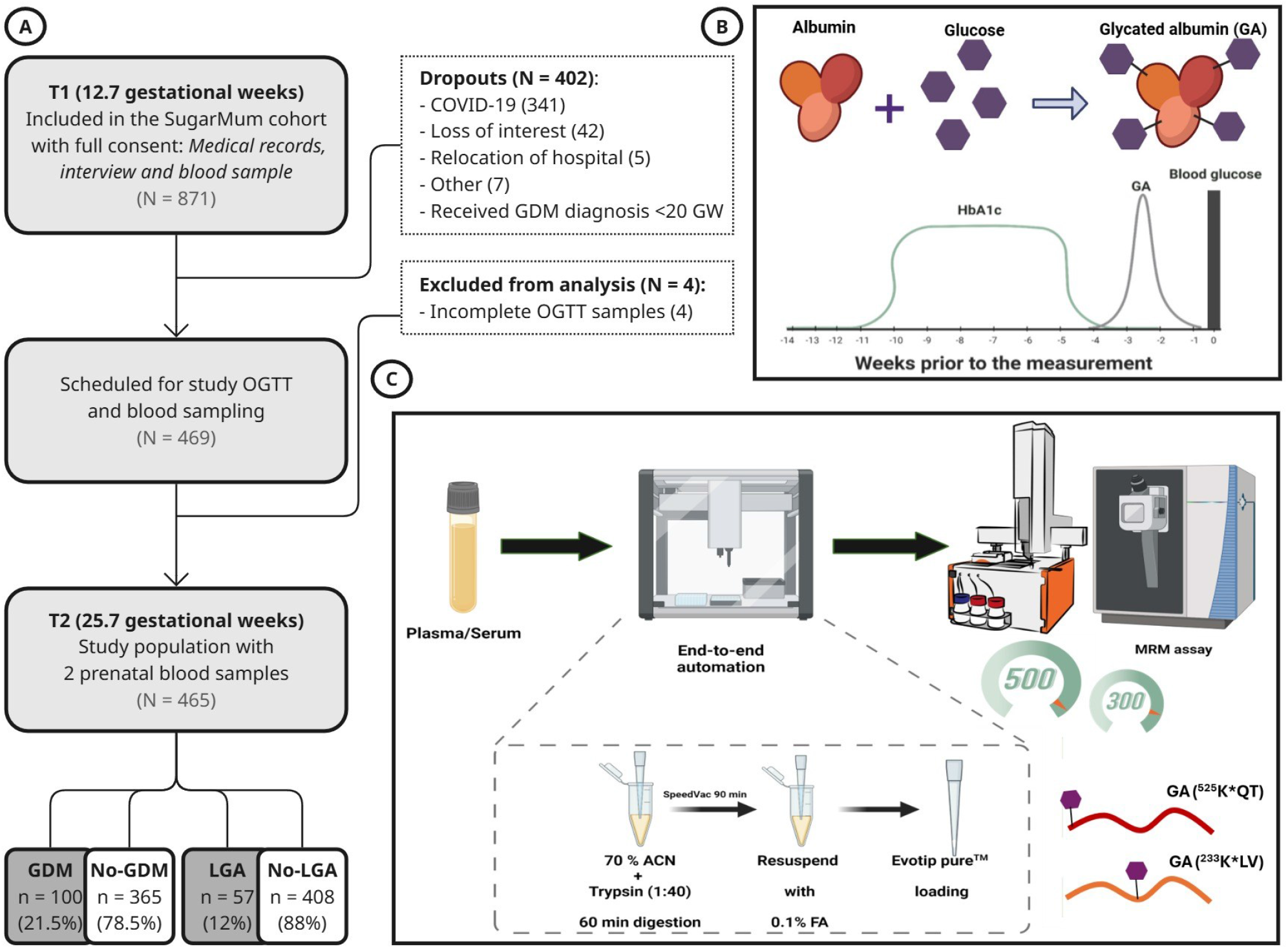
Study design, cohort and glycated albumin (GA) assay methodology. A) Study design and participant flowchart of the Hilleroed SugarMum cohort. Data are presented as n (%) of the study cohort (N = 465). Gestational age at each visit is presented as the median weeks of gestation. B) Schematic illustration of the different time windows of glycaemic exposure reflected by glucose, GA and HbA1c at the time of blood sampling. Glucose reflects current glycaemia, whereas GA and HbA1c reflect average glycaemia over approximately the preceding 2-3 weeks and 2-3 months, respectively. C) Schematic overview of the GA assay methodology. Abbreviations: OGTT, oral glucose tolerance test; GDM, gestational diabetes mellitus; LGA, large-for-gestational-age; T1, early pregnancy visit; T2, mid-late pregnancy visit; MRM, multiple reaction monitoring; ACN, acetonitrile; FA, formic acid.

### Data Collection and Outcome Definitions

Clinical characteristics and pregnancy outcomes were obtained from personal interview at the participant’s inclusion interview and from medical records. The pre-pregnancy BMI was calculated using the height measured at inclusion and the self-reported weight before conception. A family history of diabetes was defined as having one or more first- or second-degree relatives with diabetes. All participants underwent an OGTT at T2, with plasma glucose measured at fasting, 1-hour, and 2-hour time points. GDM was defined according to the WHO2013 criteria [15]. LGA was defined as the birth weight adjusted for gestational age and sex, above the >90th percentile based on a national reference population [16]. The infant was determined to have neonatal hypoglycaemia if the plasma glucose concentration was <2.5 mmol/L within the first two hours of life. Further details on data collection, outcomes, and covariates have been reported previously [14].

### Blood Sample Collection

Non-fasting venous blood samples were obtained at T1 (mean GW 12.7) and fasting venous blood samples were obtained at T2 at the time of the OGTT (mean GW 25.7) (Fig. 1A). Venous blood samples were collected in sodium-fluoride-EDTA-citrate (FC-Mix), EDTA and serum separation tubes; the last was allowed to coagulate at room temperature for at least 30 minutes, before the serum was separated by centrifugation. Samples where either analysed immediately or aliquoted and stored in a research biobank at -80° C until further analysis.

### Glycated Haemoglobin

HbA_1c_ was measured in fresh EDTA-anticoagulated whole blood using an NGSP-certifed method by high-performance LC on a Tosoh G11 or G8 Analyzer (Tosoh Bioscience, Inc.).

### LC-MS/MS Assay for Glycated Albumin

We developed a novel method for fast measurement of GA using LC-MS/MS (Fig 1B-C). The method includes acetonitrile-assisted digestion of proteins with trypsin with further parallel quantitation of glycated and non-glycated tryptic peptides. The application of highly specific multiple-reaction-monitoring (MRM)-MS allowed the use of a rapid (300 or 500 samples per day [SPD]) liquid chromatography separation optimized on an Evosep One LC system (Evosep Biosystems). Sample preparation for LC-MS/MS was performed on an automated Opentrons pipetting robot OT-2 (Opentrons Labworks) or manually in 96-well plates.

In brief, 2.5 µL of serum is diluted in 300 µL of 50 mM Tris-formate buffer prior to transfer of 20 µL to a 96 well plate. 90 µL of 70% acetonitrile and 10 µL 0.025 µg/µL trypsin (V5111 Promega) were added to serum samples and incubated at room temperature for 70 min. The digestion was quenched by adding 20 µL of 4% formic acid (FA) and the digest was dried using a speedvac concentrator centrifuge. Dried samples were resuspended in 300 µL 0.1% FA.

A total of 20 µL of each sample was loaded onto an EvoTip according to the manufacturer’s recommended protocol (Evosep Biosystems). Peptides were separated on a reverse-phase Endurance column (EV1107; 4 cm × 150 µm, 1.9 µm; Evosep Biosystems) using an integrated Evosep One microflow LC system coupled to a TSQ Altis triple quadrupole mass spectrometer equipped with an EASY-Spray ion source (Evosep Biosystems and Thermo Fisher Scientific). Chromatographic separation was performed at 300 SPD (4.8 min cycle time) using LC-MS-grade solvents consisting of buffer A (0.1% formic acid in water) and buffer B (100% acetonitrile containing 0.1% formic acid).

Percentage of GA was measured for the most abundant glycation-site in albumin ^525^K[+162]QTALVELVK termed GA (KQT) [17, 18] and divided by the sum of the glycated and its nonglycated partially digested form ^525^KQTALVELVK. A second glycation site [GA (KLV)] was measured at AEFAEVS^233^K[+162]LVTDLTK [19, 20] and related to the sum of the glycated and unglycated peptide ^226^AEFAEVSKLVTDLTK. In addition, we included peptide ^524^QTALVELVK as a digestion control (ESM Fig. 1A). Three transitions per peptide were analysed and all MS conditions are summarized in ESM Table 1. GA-total refers to the sum of GA quantified by the GA (KQT) and GA (KLV) methods.

**Table 1.** Maternal clinical and biochemical characteristics and pregnancy outcomes of the study population, stratified by GDM status.

|  |  | All | GDM | Non-GDM controls | P-value<sup>b</sup> |
|---|---|---|---|---|---|
|  | n | 465 (100.0) | 100 (21.5) | 365 (78.5) |  |
| **Maternal Characteristics** |  |  |  |  |  |
| Age (years) | 465 | 31.9 (4.5) | 31.6 (4.6) | 32.0 (4.4) | 0.509 |
| BMI (kg/m<sup>2</sup>)<sup>a</sup> | 465 | 23.9 (5.7) | 26.4 (6.8) | 23.3 (4.9) | **<0.001** |
| Origin: Denmark (n, %) | 464 | 376 (81) | 82 (82) | 294 (81) | 0.886 |
| Familial DM predisposition (n, %) | 463 | 176 (38) | 41 (42) | 135 (37) | 0.413 |
| Prior GDM (n, %) | 465 | 4 (0.9) | 4 (4.0) | 0 (0) | **0.002** |
| Nulliparous (n, %) | 465 | 180 (39) | 40 (40) | 140 (38) | 0.817 |
| **Blood samples** |  |  |  |  |  |
| OGTT (23-31 GW) |  |  |  |  |  |
| Fasting glucose (mmol/L) | 465 | 4.7 (0.4) | 5.3 (0.3) | 4.5 (0.3) | **<0.001** |
| 1-h glucose (mmol/L) | 465 | 7.0 (1.7) | 8.7 (1.6) | 6.5 (1.3) | **<0.001** |
| 2-h glucose (mmol/L) | 465 | 6.0 (1.3) | 7.1 (1.4) | 5.7 (1.1) | **<0.001** |
| GW at T1 (weeks) | 434 | 12.7(0.5) | 12.8 (0.6) | 12.7 (0.5) | 0.197 |
| GW at T2 (weeks) | 463 | 25.7 (2.4) | 25.9 (2.8) | 25.6 (2.3) | 0.158 |
| T1 GA (KQT) (%) | 436 | 6.4 (0.8) | 6.2 (0.9) | 6.4 (0.8) | **0.007** |
| T2 GA (KQT) (%) | 455 | 5.9 (0.7) | 5.8 (0.8) | 6.0 (0.7) | **0.036** |
| T1 GA (KLV) (%) | 436 | 1.4 (0.1) | 1.3 (0.2) | 1.4 (0.1) | **<0.001** |
| T2 GA (KLV) (%) | 455 | 1.3 (0.1) | 1.2 (0.2) | 1.3 (0.1) | 0.616 |
| T1 GA-total (%) | 436 | 7.7 (0.9) | 7.5 (1.0) | 7.8 (0.9) | **0.004** |
| T2 GA-total (%) | 455 | 7.2 (0.8) | 7.0 (0.9) | 7.2 (0.8) | 0.058 |
| T1 HbA<sub>1c</sub> (mmol/mol) | 435 | 31.9 (2.7) | 32.8 (2.4) | 31.6 (2.7) | **<0.001** |
| HbA<sub>1c</sub> (%) |  | 5.1 (0.2) | 5.2 (0.2) | 5.0 (0.2) |  |
| T2 HbA<sub>1c</sub> (mmol/mol)<sup>a</sup> | 464 | 30.0 (3.0) | 31.0 (4.0) | 30.0 (4.0) | **<0.001** |
| HbA<sub>1c</sub> (%)<sup>a</sup> |  | 4.9 (0.3) | 5.0 (0.4) | 4.9 (0.4) |  |
| **Pregnancy outcomes** |  |  |  |  |  |
| GW at birth (weeks) | 463 | 40.3 (1.9) | 39.7 (2.0) | 40.3 (1.7) | **<0.001** |
| Caesarean section (n, %) | 463 | 97 (21) | 29 (29) | 68 (19) | **0.037** |
| Preeclampsia (n, %) | 463 | 19 (4.1) | 7 (7.0) | 12 (3.3) | 0.148 |
| Neonatal birth weight (kg) | 459 | 3.6 (0.5) | 3.7 (0.5) | 3.6 (0.5) | 0.080 |
| z-scored neonatal birth weight | 459 | 0.1 (1.0) | 0.4 (1.1) | 0.0 (1.0) | **<0.001** |
| LGA (n, %) | 459 | 57 (12) | 21 (21) | 36 (10) | **0.005** |
| Neonatal hypoglycaemia (n, %) | 459 | 17 (3.7) | 10 (10) | 7 (1.9) | **<0.001** |
| Neonatal intensive care admission (n, %) | 459 | 36 (7.8) | 15 (15) | 21 (5.8) | **0.005** |

Data are presented as n (%) for categorical variables and mean (SD) or median (IQR) for continuous variables. T1 and T2 refer to measurements obtained from blood samples collected in early pregnancy and mid-late pregnancy, respectively.
<sup>a</sup>Data are presented as median (IQR) for non-normally distributed variables.
<sup>b</sup>Categorical variables were compared using Fisher’s exact test, normally distributed continuous variables using Welch’s Two Sample t-test, and skewed variables using the Mann-Whitney U test.
Statistically significant p-values (<0.05) are shown in bold.

Single point calibration was performed using human serum Seronorm™ Immunoassay Liq L-2 (SERO) whose GA% was determined according to the primary calibrator; JCCRM-611 certified GA standard (ReCCS, Japan) at three levels (M, H, HH). Seronorm™ Immunoassay Liq L-1 serum pool was used as quality control (QC) and distributed evenly in quintuplicates across each 96-well plate. Mean inter-assay CV was 6.2% and 6.1% for GA (KQT) and GA (KLV), respectively (ESM Fig. 1B-C). All MRM-MS raw files were processed in Skyline and further processed in Jupyter Notebook and all statistical calculations were performed in RStudio (RStudio Team [2020]).

Total GA was measured in plasma by an FDA-approved enzymatic assay with Glycated Albumin-L reagent kits (Asahi Kasei Pharma, Tokyo, Japan) on a cobas 6000 analyser (Roche Diagnostics) as previously described [21]. The inter-assay coefficients of variation for GA were 1.7% at 0.58 g/L and 4.8% at 1.67 g/L, respectively. GA is expressed as a percentage of total albumin, which was measured with bromocresol purple on a cobas 6000 analyser.

### Statistical Analyses

All statistical analyses were performed in R version 4.5.3. Baseline characteristics for GDM and LGA case participants, and their respective control group were compared using Fisher’s exact test for categorical variables, Welch’s Two Sample t-test for normally distributed and continuous variables, and Mann-Whitney U test for skewed, but continuous variables.

Prospective associations of GA and HbA_1c_ measured at T1 and T2 with the risk of GDM and LGA were assessed using logistic regression models. All models were adjusted for pre-gestational BMI and maternal age at delivery. Biomarker levels (GA (KQT), GA (KLV), GA-total, and HbA_1c_) were standardized to z-scores separately at each pregnancy time point. Odds ratios (ORs) are reported per one SD increase. Observations with an absolute z-score >5 were considered extreme outliers. One extreme outlier was identified among the early pregnancy HbA_1c_ measurements and was therefore excluded from the respective analyses.

Differences in GA levels between LGA and non-LGA controls and between GDM and non-GDM controls were assessed using linear mixed-effects regression models. Models included outcome status (LGA or GDM), study visit, and their interaction as fixed effects and were adjusted for family history of diabetes (binary), nulliparity (binary), pre-gestational BMI (continuous), maternal age at delivery (continuous), and gestational age at blood collection (continuous). Participant ID was included as a random intercept to account for repeated measurements within individuals. Estimated marginal means were used to obtain adjusted comparisons between outcome groups at each study visit. GA levels are presented as raw means with 95% confidence intervals (CIs). The packages *lme4* and *emmeans* were applied. Spearman correlation analyses were performed to assess the relationship between maternal pre-gestational BMI and each biomarker (GA (KQT), GA (KLV), GA-total, and HbA_1c_).

## Results

### Cohort characteristics

Maternal age, Danish origin, family history of diabetes, and parity were comparable between women with GDM and non-GDM controls. However, women with GDM had a higher pre-pregnancy BMI and were more likely to have a previous history of GDM. GDM was also associated with a higher frequency of preterm delivery, caesarean section, and LGA, as well as an increased incidence of neonatal hypoglycaemia and NICU admission (Table 1). Compared with non-LGA controls, women in the LGA group were more likely to be of Danish origin (93% vs 79%) and had higher fasting and 1-hour OGTT glucose concentrations. Their neonates had higher birth weight and were more likely to experience neonatal hypoglycaemia and require NICU admission (ESM Table 2).

**Table 2.** Plasma GA (KQT), GA (KLV) and GA-total means in early and mid-late pregnancy by LGA status.

|  | **LGA** |  | **Non-LGA controls** |  |  |
|---|---|---|---|---|---|
|  | *Mean* | *95% CI* | *Mean* | *95% CI* | *P-value<sup>a</sup>* |
| **T1 (N = 430)** |  |  |  |  |  |
| GA (KQT) (%) | 6.51 | 5.19-7.82 | 6.35 | 4.63-8.03 | **0.027** |
| GA (KLV) (%) | 1.36 | 1.06-1.65 | 1.35 | 1.07-1.59 | 0.230 |
| GA-total (%) | 7.87 | 6.26-9.32 | 7.71 | 5.74-9.53 | **0.032** |
| **T2 (N = 449)** |  |  |  |  |  |
| GA (KQT) (%) | 6.06 | 4.97-7.48 | 5.90 | 4.66-7.39 | **0.048** |
| GA (KLV) (%) | 1.29 | 1.05-1.47 | 1.25 | 1.01-1.50 | **0.010** |
| GA-total (%) | 7.35 | 6.04-8.92 | 7.15 | 5.66-8.82 | **0.030** |

GA measurements are presented as the mean (95% CI).
<sup>a</sup>Differences of GA means between LGA case and non-LGA control participants at each visit were assessed using linear mixed-effect regression models adjusted for family history of diabetes, nulliparity, pre-gestational BMI, maternal age at delivery, and gestational age at blood sampling with a participant-specific random intercept.

### Comparisons of GA and HbA_1c_ between GDM and Control Participants

GA levels were measured by the developed LC-MS/MS method (Fig. 1C). All GA levels were lower at T1 in women with GDM compared to controls (GA-total; 7.5% (1.0) vs. 7.8% (0.8), *p*=0.004). At T2 all GA levels tended to be lower in the GDM group, but only significant for GA (KQT) (5.8% (0.8) vs. 6.0 (0.7), *p*=0.036). Participants with GDM had higher levels of HbA_1c_ at T1 with mean (SD) 32.8 mmol/mol (2.4) for GDM participants vs. 31.6 mmol/mol (2.7) for non-GDM controls (*p*<0.001). HbA_1c_ levels were also higher for GDM participants at T2 with a median (IQR) of 31.0 mmol/mol (4.0) for the GDM women vs. 30.0 mmol/mol (4.0) for the control group (*p*<0.001) (Fig. 2 and Table 1).

**Fig. 2.**
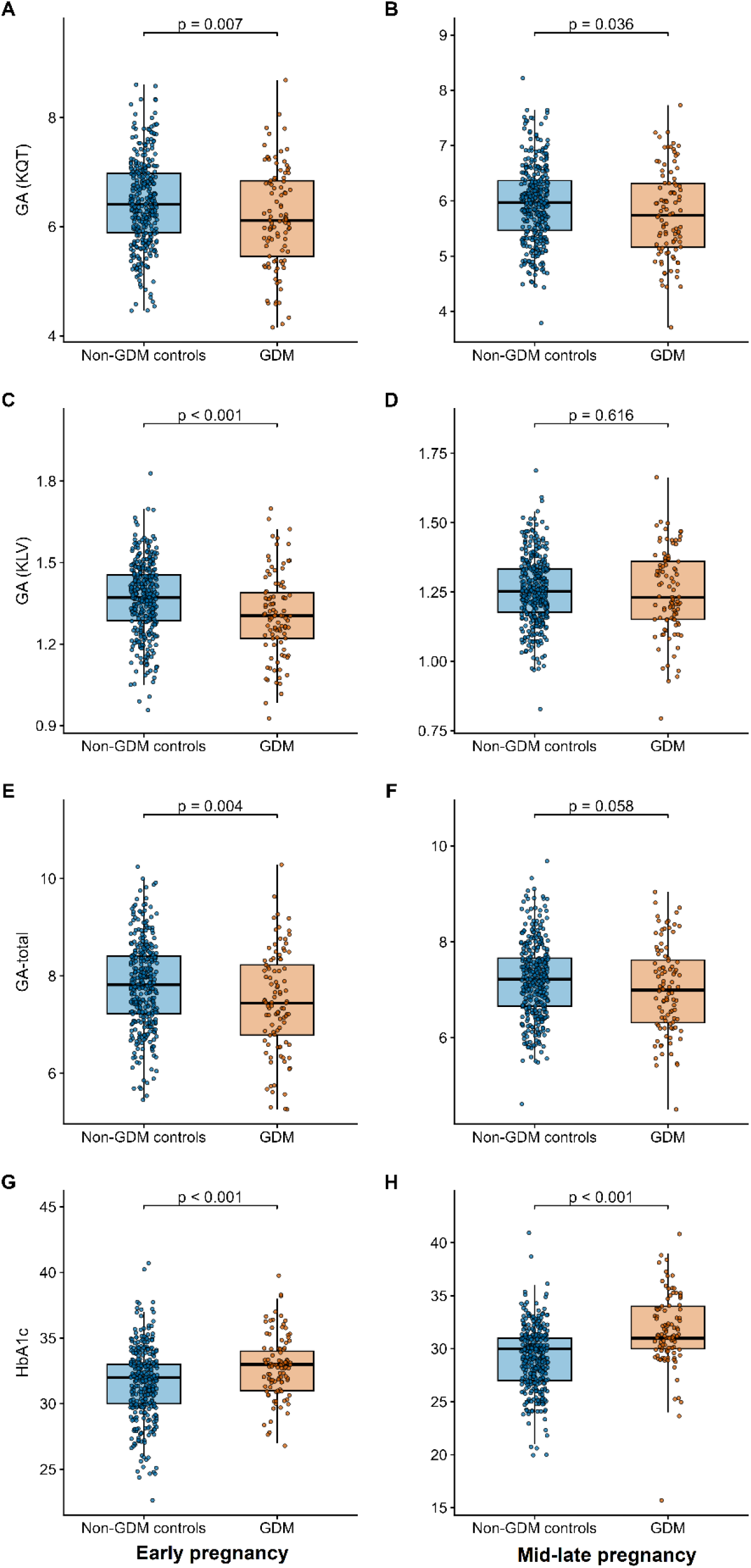
Boxplots showing the distributions of (A, B) GA (KQT), (C, D) GA (KLV), (E, F) GA-total and (G, H) HbA1c in early (left) and mid-late (right) pregnancy between GDM case (orange) and non-GDM controls (blue). Between-group differences were assessed using the Wilcoxon rank-sum test for mid-late pregnancy HbA1c, owing to its non-normal distribution, and the two-sample t test for all other measures.

### Comparisons of GA and HbA_1c_ between pregnancies with and without LGA

GA (KLV) was higher at T2 in the LGA group than in the non-LGA control group (1.28% (0.1) vs. 1.25% (0.1), *p*=0.027). All other measures of GA and HbA_1c_ levels also tended to be higher in the LGA group than in the non-LGA control group, but none of these differences were significant (ESM Fig. 2 and ESM Table 2).

### Prospective Associations of GA, HbA_1c_ and the Risk of GDM and Adverse Pregnancy Outcomes

Lower GA was not associated with a higher risk of GDM at either T1 or T2, as estimated by ORs (95% CI). In contrast, HbA_1c_ was positively associated with subsequent development of GDM at both T1 and T2 (*p*<0.001). The corresponding ORs (95% CI) were 2.20 (1.46-3.38) and 2.30 (1.75-3.09), respectively (Fig. 3A-B). GA was significantly associated with the risk of LGA at both T1 and T2. At T1, higher GA-total and GA (KQT) were significantly associated with higher odds of LGA, with ORs (95% CI) of 1.59 (1.13-2.27) and 1.59 (1.13-2.25), respectively. At T2, all three GA exposures were significantly associated with LGA, with ORs ranging from 1.42 to 1.51. Conversely, HbA_1c_ was not associated with the risk of LGA at either time point (Fig. 3C-D).

**Fig. 3.**
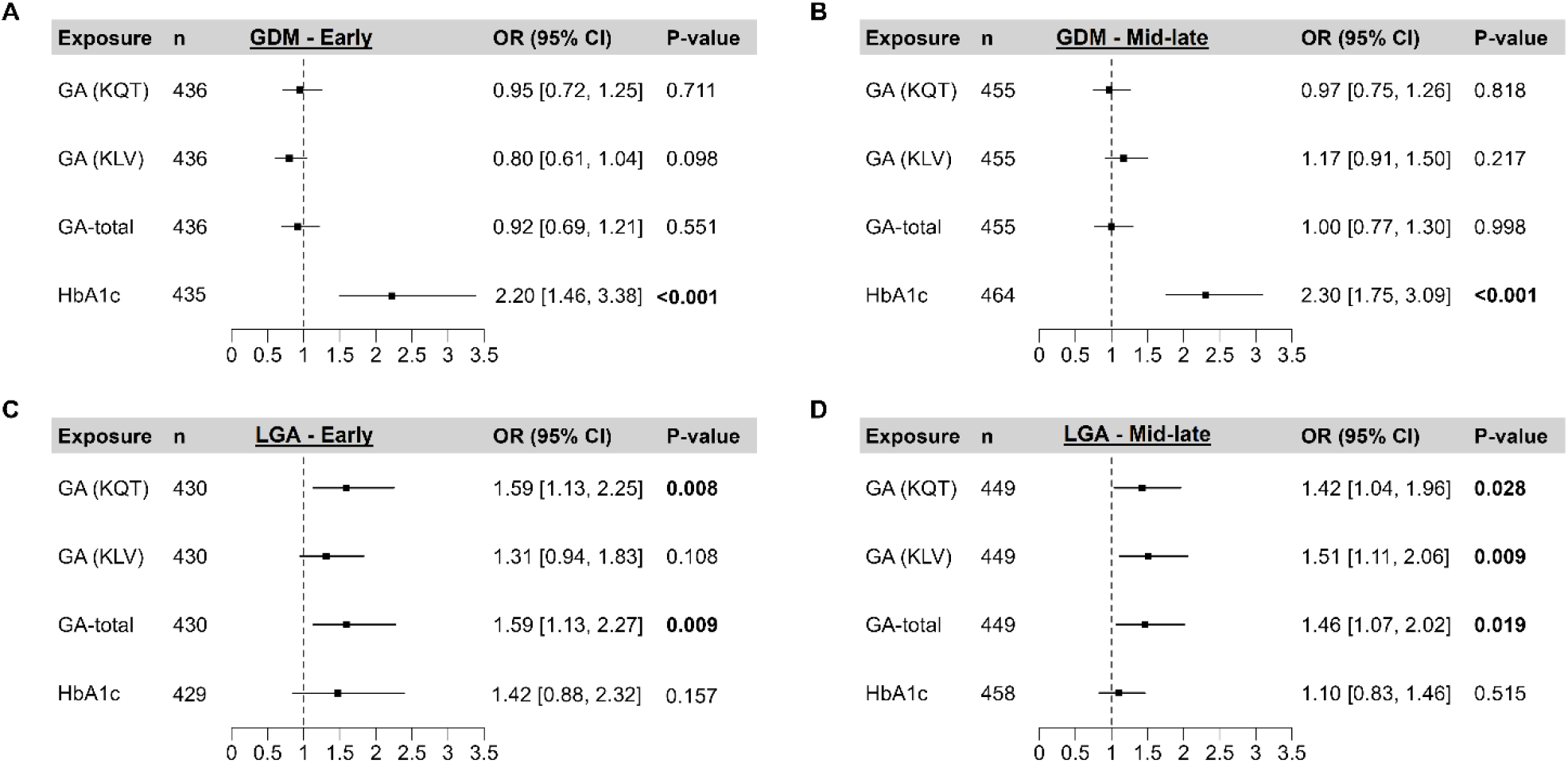
Forest plots of odds ratios (ORs) (95% CI) obtained from logistic regression models adjusted for maternal age at delivery and pre-pregnancy BMI. ORs represent the change in odds of the outcome associated with a one standard deviation increase in the z-scored exposure. (A) Outcome: GDM; exposures measured in early pregnancy. (B) Outcome: GDM; exposures measured in mid-late pregnancy. (C) Outcome: LGA; exposures measured in early pregnancy. (D) Outcome: LGA; exposures measured in mid-late pregnancy. The x-axis shows ORs. The vertical dashed line indicates the reference value of no effect (OR = 1). Squares represent OR estimates and horizontal lines represent 95% CIs. Statistically significant p values (<0.05) are shown in bold.

In the unadjusted OR analyses, GA was significantly associated with GDM for all measures except GA (KLV) at T2. In contrast, no significant associations with LGA were observed before adjustment, except for GA (KLV) at T2 (ESM Table 3).

We also investigated associations between GA, HbA_1c_ and the risk of caesarean section, neonatal hypoglycaemia, NICU admission, and a composite neonatal adverse outcome comprising shoulder dystocia, nerve injury and bone fracture. GA was not significantly associated with any of these outcomes. HbA_1c_ was associated with caesarean section and neonatal hypoglycaemia at T2, with adjusted ORs (95% CI) of 1.28 (1.01-1.62) and 1.87 (1.15-3.10), respectively (ESM Table 4).

### Association between GA and risk of LGA is most pronounced at low GA z-scores

We analysed adjusted associations between biomarker levels at T1 and T2 and the odds of LGA. At T1, GA (KQT) and GA-total were significantly and positively associated with LGA, whereas GA (KLV) showed no significant association (Fig. 4A-C). At T2, all GA exposures were positively and significantly associated with LGA (Fig. 4E-G), indicating that lower GA z-scores were consistently associated with lower odds of LGA. The precision of the estimates is greatest at lower z-scores, where the confidence intervals are narrower. HbA_1c_ showed a positive but non-significant association with the odds of LGA at both T1 and T2, (Fig. 4D and Fig. 4H).

**Fig. 4.**
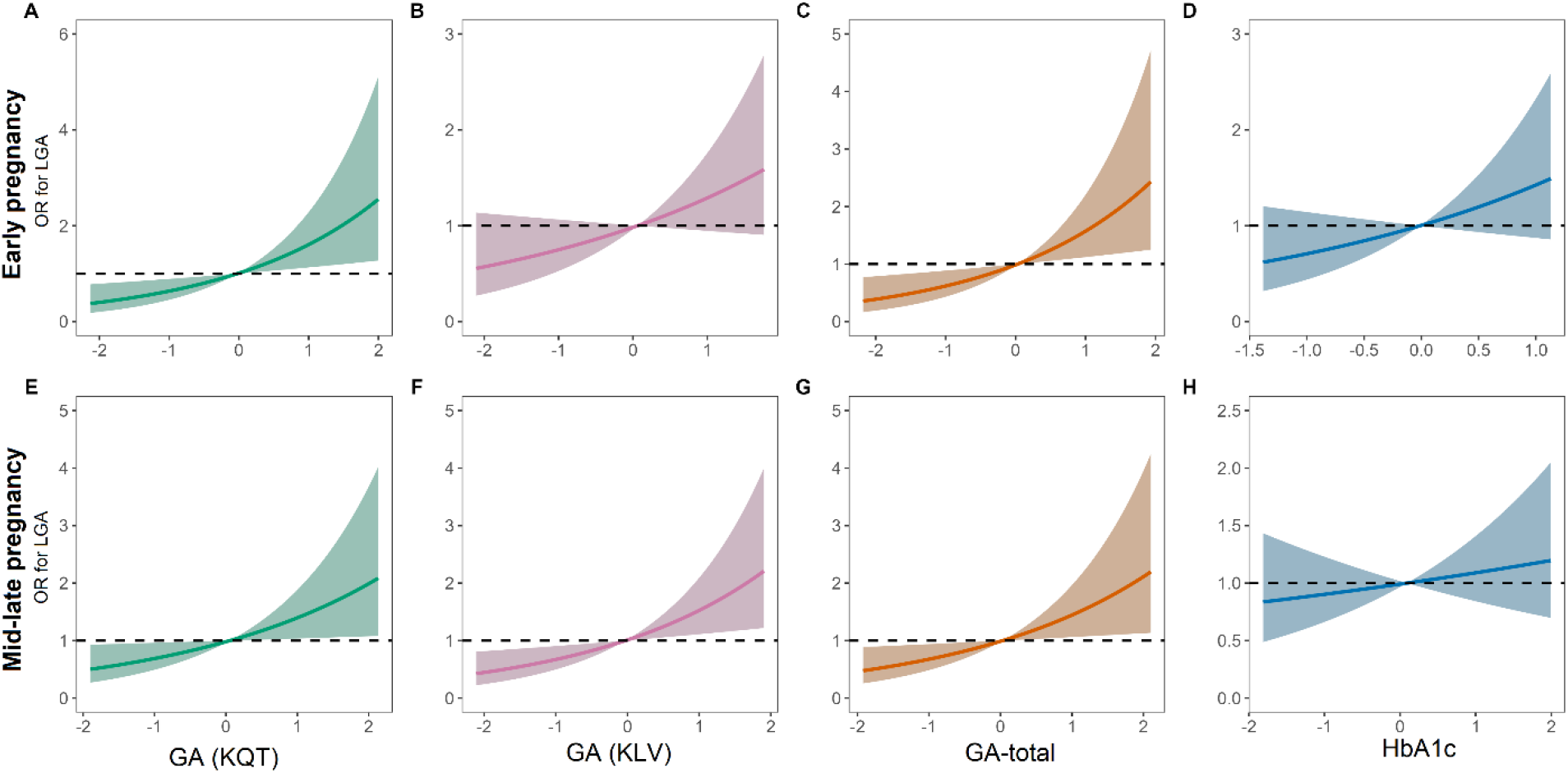
Associations of (A, E) GA (KQT), (B, F) GA (KLV), (C, G) GA-total, and (D, H) HbA1c with OR for LGA, estimated from logistic regression models adjusted for maternal age at birth and pre-pregnancy BMI. Panels show the association between z-scored biomarker concentrations and ORs (95% CI) for LGA. Top panels correspond to early pregnancy, and bottom panels to mid-late pregnancy. Y-axes represent ORs for LGA. The horizontal dashed line indicates the reference value of no effect (OR = 1).

### Linear mixed-effects regression models for GA by LGA status

Linear mixed-effects models showed significantly higher GA levels in women who delivered an LGA infant across all GA measures at both timepoints, with the exception of GA (KLV) at T1 (Table 2). In contrast, no significant differences in GA levels were observed according to GDM status using the same modelling approach (ESM Table 5).

### GA, HbA_1c_ and pre-gestational maternal BMI Correlation Analysis

Spearman correlation analyses demonstrated that all GA measures showed a strong inverse correlation with pre-gestational BMI at both time points (T1 and T2; all *p*<0.001) (Fig. 5A-C and Fig. 5E-G). In contrast, HbA_1c_ showed a weak positive correlation with pre-gestational BMI at T2 (R=0.21, *p*<0.001), but no significant correlation at T1 (*p*=0.59) (Fig. 5D and Fig. 5H).

**Fig. 5.**
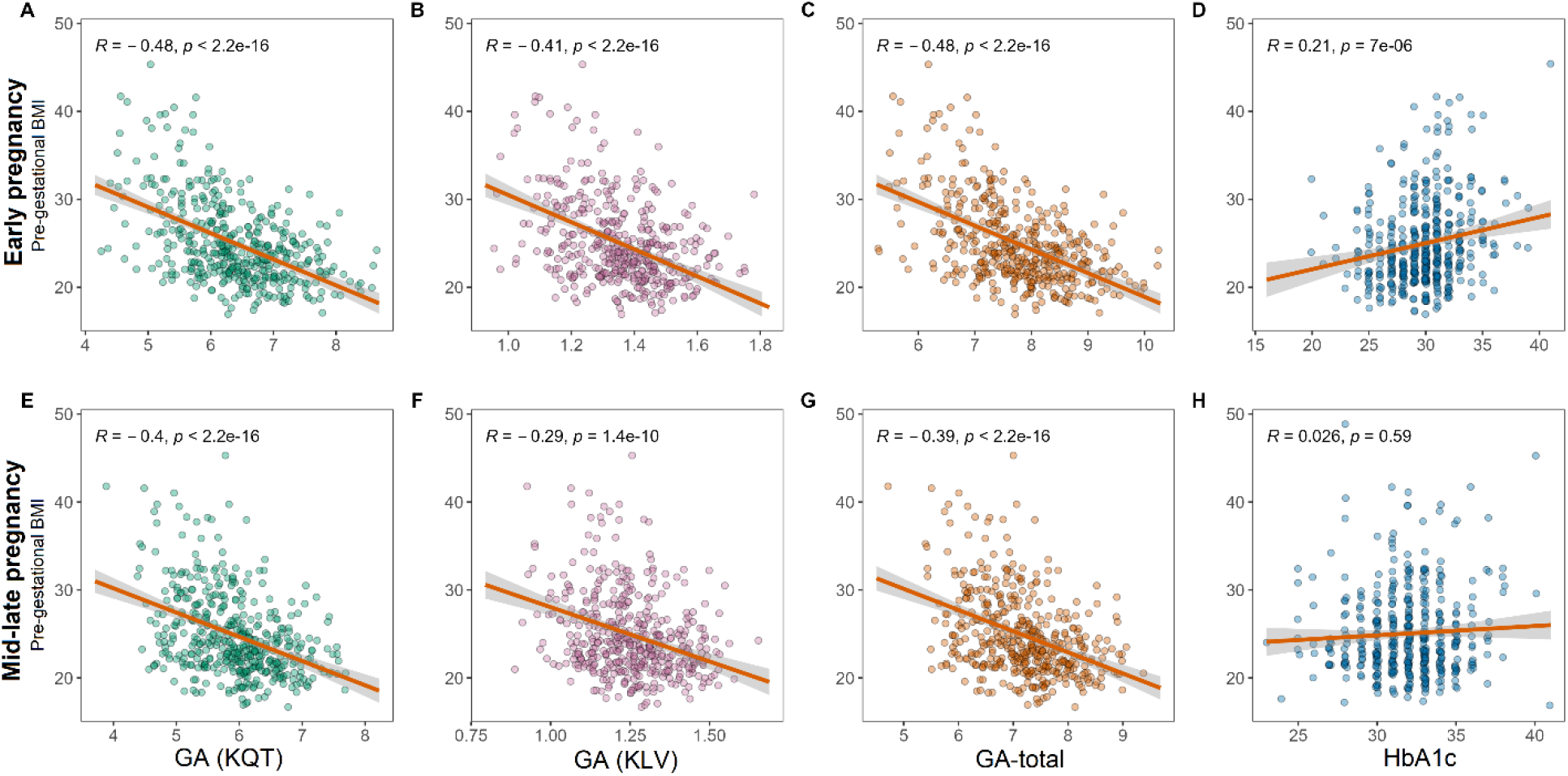
Association of maternal pre-gestational BMI with GA and HbA1c. Linear regression models showing the associations of maternal pre-gestational BMI with (A, E) GA (KQT), (B, F) GA (KLV), (C, G) GA-total, and (D, H) HbA1c. Top panels (A-D) correspond to early pregnancy, and bottom panels (E-H) correspond to mid-late pregnancy. Y-axes represent pre-gestational BMI. Regression lines and 95% confidence intervals were estimated using ordinary least squares linear regression. Spearman’s rank correlation coefficient (R) and corresponding P-values are shown in each panel.

## Discussion

In this prospective longitudinal cohort study, elevated GA was associated with LGA, but not with GDM. In contrast, elevated HbA_1c_ was associated with GDM but showed no association with LGA. These findings were consistent at both T1 and T2, suggesting that the two glycaemic markers may capture different aspects of conditions associated with maternal glycaemic physiology during pregnancy. However, whether GA provides additional information on LGA risk beyond HbA_1c_ cannot be determined from the present study and requires confirmation in larger, independent cohorts before GA can be considered a clinically useful biomarker of LGA risk.

To our knowledge, this is the first prospective longitudinal cohort study to demonstrate an association between higher maternal GA and the subsequent delivery of an LGA infant. Previous studies have primarily evaluated GA in women with established GDM, measured GA late in pregnancy or after delivery, or found no significant association with LGA in unselected populations [22–24]. Our findings suggest that GA may reflect metabolic processes relevant to fetal overgrowth that are not fully captured by HbA_1c_ or by the diagnosis of GDM alone. This may be particularly relevant given that fetal overgrowth can occur in the absence of clinically diagnosed GDM (Table 1).

Higher HbA_1c_ levels in early pregnancy have previously been associated with subsequent development of GDM [25] and adverse fetal growth outcomes, including LGA [26]. In line with these findings, we observed an association between HbA_1c_ and GDM at both T1 and T2. However, while a positive trend was observed for LGA, the association was not statistically significant in our study.

While conflicting results have been reported regarding the prospective association between GA and GDM [22, 27–31], our findings are consistent with those of a previous prospective study in U.S. women, which observed significantly lower GA concentrations at 15-26 GW among participants who subsequently developed GDM (*p*=0.004) [32]. Notably, studies reporting higher GA levels among women who subsequently develop GDM have primarily been conducted in Asian populations. Whether these differences reflect ethnic variation, differences in statistical power, or a combination of both remains unclear. Although the GA assay methodologies differed between our study and the U.S. study, method comparison analyses showed that GA (KQT), GA (KLV), and GA-total were all significantly correlated with the enzymatic Lucica GA-L assay used in the study by Cole and colleagues [32, 33] (ESM Fig. 1D-F), despite differences in the GA% ranges measured by the respective methods (ESM Fig. 1A).

The observation that GA was lower in women who developed GDM may seem counterintuitive, given that GA reflects glycaemic exposure (Fig. 2 and Table 1). Although GA is known to decrease throughout pregnancy, this physiological change does not explain the lower GA levels observed in women who developed GDM [34]. We and others [34, 35] have reported a significant inverse association between GA and pre-gestational BMI (Fig. 5), suggesting that BMI may partly explain this finding. The importance of accounting for pre-gestational BMI was further underscored by its substantial impact on the associations between GA and both GDM and LGA. Adjustment for BMI attenuated the associations between GA and GDM, whereas significant associations between GA and LGA emerged only after adjustment, with the exception of GA (KLV) at T1, for which the association remained non-significant (ESM Table 3). These findings suggest that pre-gestational BMI is an important potential confounder in the relationship between GA and pregnancy outcomes and may partly account for the observed associations.

In this study we measured albumin glycation at two distinct sites (KQT and KLV). GA (KQT) and GA-total were significantly associated with LGA in early pregnancy, whereas GA (KLV) was not. The association observed for GA-total therefore appears to be driven primarily by the GA (KQT) component. These findings suggest that GA (KQT) may be particularly relevant for assessing LGA risk in early pregnancy. Further secondary analysis showed no significant associations between GA and other pregnancy outcomes. In contrast, HbA_1c_ was associated with both neonatal hypoglycaemia and caesarean section at T2. ORs were also estimated for NICU admission and a composite neonatal adverse outcome. However, none of these investigated associations reached statistical significance for either biomarker.

Taken together, these findings suggest that HbA_1c_ and GA may capture distinct aspects of metabolic status that are differentially associated with pregnancy and neonatal outcomes.

Strengths and limitations apply to this study; The SugarMum cohort was designed to evaluate the WHO2013 diagnostic criteria for GDM in a prospective unselected Danish cohort. The strengths of this study include its population-based design, which enhances generalisability and reduces the potential for selection bias related to socioeconomic status and differences in comorbidity burden. Data from eligible women, confirmed that the study participants were comparable to non-participants on most baseline characteristics [14]. Blood samples were drawn at enrolment at mean 12.7 GW and at OGTT mean 25.7 GW and allowed for prospective assessment of associations of GA with GDM and LGA risk.

Regarding assay methodology, strengths included high-specificity LC-MS/MS analysis of two albumin glycation sites using a novel assay with high assay precision (ESM Fig. 1B-C). The assay enabled site-specific assessment of albumin glycation at two distinct sites: the highly glycation-sensitive KQT site and the less glycated KLV site (ESM Fig. 1A). Albumin has multiple glycation sites; however, our assay was designed to capture the most abundant glycations [17–20].

Although method comparison analyses demonstrated a strong correlation between these sites (ESM Fig. 1D-E), the strongest and most consistent associations with study outcomes were observed for GA (KQT) and GA-total. GA measurements were performed on stored biobank samples, but consistent sample handling and storage procedures in this cohort minimise the risk of preanalytical variability. Nevertheless, the potential effects of long-term storage on GA stability cannot be completely excluded and could be investigated in future studies.

In conclusion, higher GA at T1 and T2 were associated with increased LGA risk. In contrast, HbA_1c_ was not significantly associated with LGA risk at either timepoint. These findings suggest that GA may capture aspects of maternal metabolism relevant to fetal overgrowth that are not reflected by HbA_1c_. In contrast, GA was not associated with GDM risk at either timepoint, whereas HbA_1c_ was positively associated with GDM risk at both T1 and T2.

## Supporting information

Electronic supplementary material

## Acknowledgments

We would like to thank the women participating in the SugarMum study for their participation and Amalie Bøgsted and Maureen Sampson for excellent technical assistance in performing the laboratory analysis.

## Data availability

Data are available from the corresponding author on reasonable request.

## Authors relationship and activities

The authors declare no conflict of interest.

The contributions of the NIH author is considered Works of the United States Government. The findings and conclusions presented in this paper are those of the author(s) and do not necessarily reflect the views of the NIH or the U.S. Department of Health and Human Services.

## Contribution statement

CFJ, TSC and MO conducted data analysis, interpretation and drafting of the manuscript. CS, DMJ, HDM, PD, KH, MO and TDC designed the study and contributed to funding applications. CS, TDC, LR, CE and MO obtained legal approvals. BR, DBS, DBN, NB, OV and MO contributed to the development of the GA assay. All authors contributed to data analysis, interpretation and critical revision of the manuscript and approved the final version for publication. MO is the guarantor of this work and, as such, has full access to all the data in the study and takes responsibility for the integrity of the data and the accuracy of the data analysis.

## Abbreviations

GA: Glycated albumin
GA (KLV): GA measured at site 233-KLV
GA (KQT): GA measured at site 525-KQT
GA-total: Sum of GA (KQT) and GA (KLV)
GDM: Gestational diabetes mellitus
GW: Gestational weeks
HbA_1c_: Haemoglobin A_1c_
LGA: Large-for-gestational-age
OGTT: Oral glucose tolerance test
OR: Odds ratio
T1: Early pregnancy
T2: Mid-late pregnancy

