## Supplementary material for "Elevated Maternal Glycated Albumin is Associated with Large for Gestational Age Infants: Results from a Prospective Longitudinal Study": Electronic supplementary material

**Electronic supplementary material (ESM)**

**Carolina F. Jørgensen et al.**

**Supplementary Tables 1-5 and Figure 1-2**

**Supplementary Table 1**. Albumin MRM assay peptides.

| **Target peptides (tryptic)** *P02768, Albumin* | **Position in amino acid sequence** | **Precursor ion Q1 m/z (+2)** | **Fragment ion 1 m/z (+1)** | **Fragment ion 2 m/z (+1)** | **Fragment ion 3 m/z (+1)** | **CE, eV** |
|---|---|---|---|---|---|---|
| K[+162]QTALVELVK | 525 | 645.88 | y8, 872.54 | y7, 771.50 | y3, 359.27 | 26.48 |
| KQTALVELVK | 525 | 564.85 | y8, 872.54 | y7, 771.50 | y3, 359.27 | 22.22 |
| QTALVELVK | 526 | 500.81 | y7, 771.50 | y5, 587.38 | y4, 488.31 | 18.85 |
| AEFAEVSK[+162]LVTDLTK | 226 (233) | 906.98 | y3, 361.24 | y2, 248.16 | b3, 348.16 | 40.22 |
| AEFAEVSKLVTDLTK | 226 | 825.95 | y3, 361.24 | y2, 248.16 | b3, 348.16 | 35.96 |

**Supplementary Table 2.** Maternal clinical and biochemical characteristics and pregnancy outcomes of the study population, stratified by LGA status.

| | | **All** | **LGA** | **Non-LGA controls** | **P-value**[b] |
|---|---|---|---|---|---|
| | n | 465 (100.0) | 57 (12.2) | 408 (87.7) | |
| **Maternal Characteristics** | | | | | |
| Age (years) | 459 | 31.9 (4.5) | 31.4 (4.5) | 31.9 (4.4) | 0.397 |
| BMI (kg/m$^2$)[a] | 459 | 23.9 (5.7) | 25.2 (8.8) | 23.8 5.4) | 0.252 |
| Origin: Denmark (n, %) | 458 | 371 (81) | 53 (93) | 318 (79) | **0.011** |
| Familial DM predisposition (n, %) | 457 | 175 (38) | 17 (30) | 158 (40) | 0.190 |
| Prior GDM (n, %) | 459 | 4 (0.9) | 0 (0) | 4 (1.0) | >0.999 |
| Nulliparous (n, %) | 459 | 176 (38) | 16 (28) | 160 (40) | 0.109 |
| **Blood samples** | | | | | |
| OGTT (23-31 GW) | | | | | |
| Fasting glucose (mmol/L) | 459 | 4.7 (0.4) | 4.8 (0.5) | 4.6 (0.4) | **0.006** |
| 1-h glucose (mmol/L) | 459 | 7.0 (1.7) | 7.7 (1.6) | 6.9 (1.6) | **<0.001** |
| 2-h glucose (mmol/L) | 459 | 6.0 (1.3) | 6.3 (1.3) | 6.0 (1.3) | 0.051 |
| GW at T1 (weeks) | 430 | 12.7 (0.5) | 12.7 (0.4) | 12.7 (0.5) | 0.793 |
| GW at T2 (weeks) | 459 | 25.7 (2.4) | 25.9 (1.9) | 25.7 (1.4) | 0.877 |
| T1 GA (KQT) (%) | 430 | 6.4 (0.8) | 6.5 (0.8) | 6.4 (0.8) | 0.204 |
| T2 GA (KQT) (%) | 449 | 5.9 (0.7) | 6.1 (0.6) | 5.9 (0.7) | 0.090 |
| T1 GA (KLV) (%) | 430 | 1.35 (0.1) | 1.36 (0.1) | 1.35 (0.1) | 0.598 |
| T2 GA (KLV) (%) | 449 | 1.25 (0.1) | 1.28 (0.1) | 1.25 (0.1) | **0.027** |
| T1 GA-total (%) | 430 | 7.7 (0.9) | 7.9 (0.9) | 7.7 (0.9) | 0.225 |
| T2 GA-total (%) | 449 | 7.2 (0.8) | 7.4 (0.7) | 7.2 (0.8) | 0.063 |
| T1 HbA$_{1c}$ (mmol/mol) | 429 | 31.9 (2.7) | 32.4 (2.6) | 31.8 (2.7) | 0.158 |
| HbA$_{1c}$ (%) | | 5.1 (0.2) | 5.1 (0.2) | 5.1 (0.2) | |
| T2 HbA$_{1c}$ (mmol/mol)[a] | 458 | 30.0 (3.0) | 30.0 (2.0) | 30.0 (2.0) | 0.545 |
| HbA$_{1c}$ (%)[a] | | 4.9 (0.3) | 4.9 (0.2) | 4.9 (0.2) | |
| **Pregnancy outcomes** | | | | | |
| Gestational age at birth (weeks) | 459 | 40.3 (1.9) | 40.1 (1.9) | 40.3 (1.9) | 0.946 |
| C-section (n, %) | 459 | 95 (21) | 14 (25) | 81 (20) | 0.484 |
| Birth weight (kg) | 459 | 3.6 (0.5) | 4.4 (0.3) | 3.5 (0.4) | **<0.001** |
| z-scored birth weight | 459 | 0.1 (1.0) | 1.8 (0.5) | -0.2 (0.8) | **<0.001** |
| Preeclampsia (n, %) | 459 | 18 (3.9) | 2 (3.5) | 16 (4.0) | >0.999 |
| Neonatal hypoglycaemia (n, %) | 459 | 17 (3.7) | 6 (11) | 11 (2.7) | **0.012** |
| Neonatal intensive care admission (n, %) | 459 | 36 (7.8) | 9 (16) | 27 (6.7) | **0.030** |

Data are presented as n (%) for categorical variables and mean (SD) or median (IQR) for continuous variables. T1 and T2 refer to measurements obtained from blood samples collected in early pregnancy and mid-late pregnancy, respectively.

[a]Data are presented as median (IQR) for non-normally distributed variables.

[b]Categorical variables were compared using Fisher's exact test, normally distributed continuous variables using Welch's Two Sample t-test, and skewed variables using the Mann-Whitney U test.

Statistically significant *p*-values (<0.05) are shown in bold.

**Supplementary Table 3.** Crude and adjusted odds ratios for GDM and LGA associated with GA (KQT), GA (KLV) and GA-total at T1 and T2.

| | *Timepoint* | *n* | **Crude** *OR (95% CI)* | *p-value* | **Adjusted** *OR (95% CI)* | *p-value* |
|---|---|---|---|---|---|---|
| **GDM** | | | | | | |
| GA (KQT) | T1 | 436 | 0.71 (0.56-0.89) | **0.004** | 0.95 (0.72-1.25) | 0.711 |
| GA (KQT) | T2 | 455 | 0.77 (0.61-0.96) | **0.023** | 0.97 (0.75-1.26) | 0.818 |
| GA (KLV) | T1 | 436 | 0.64 (0.51-0.81) | **<0.001** | 0.80 (0.61-1.04) | 0.098 |
| GA (KLV) | T2 | 455 | 0.94 (0.75-1.17) | 0.568 | 1.17 (0.91-1.50) | 0.217 |
| GA-total | T1 | 436 | 0.69 (0.54-0.87) | **0.002** | 0.92 (0.69-1.21) | 0.551 |
| GA-total | T2 | 455 | 0.78 (0.62-0.98) | **0.038** | 1.00 (0.77-1.30) | 0.998 |
| **LGA** | | | | | | |
| GA (KQT) | T1 | 430 | 1.21 (0.90-1.62) | 0.201 | 1.59 (1.13-2.25) | **0.008** |
| GA (KQT) | T2 | 449 | 1.25 (0.94-1.67) | 0.126 | 1.42 (1.04-1.96) | **0.028** |
| GA (KLV) | T1 | 430 | 1.09 (0.81-1.46) | 0.581 | 1.31 (0.94-1.83) | 0.108 |
| GA (KLV) | T2 | 449 | 1.36 (1.02-1.82) | **0.037** | 1.51 (1.11-2.06) | **0.009** |
| GA-total | T1 | 430 | 1.20 (0.90-1.62) | 0.222 | 1.59 (1.13-2.27) | **0.009** |
| GA-total | T2 | 449 | 1.28 (0.96-1.70) | 0.097 | 1.46 (1.07-2.02) | **0.019** |

Values are odds ratios (95% CI). Significant *p*-values ($p<0.05$) are shown in bold. Adjusted models were adjusted for maternal age and pre-gestational BMI.

**Supplementary Table 4**. Associations of GA, $HbA_{1c}$ and the risk of adverse pregnancy outcomes

| | **GA-total** | | | | **$HbA_{1c}$** | | | |
|---|---|---|---|---|---|---|---|---|
| | *N* | *OR* | *95% CI* | *P* | *N* | *OR* | *95% CI* | *P* |
| *Caesarean section* | | | | | | | | |
| T1 | 434 | 0.96 | 0.73; 1.26 | 0.764 | 434 | 1.06 | 0.83; 1.31 | 0.894 |
| T2 | 453 | 1.02 | 0.79; 1.31 | 0.899 | 462 | 1.28 | 1.01; 1.62 | **0.040** |
| *SGA* | | | | | | | | |
| T1 | 430 | 0.81 | 0.52; 1.25 | 0.349 | 340 | 0.76 | 0.41; 1.21 | 0.361 |
| T2 | 449 | 0.67 | 0.44; 1.01 | 0.056 | 458 | 1.09 | 0.75; 1.59 | 0.661 |
| *Neonatal hypoglycaemia* | | | | | | | | |
| T1 | 430 | 1.47 | 0.82; 2.63 | 0.191 | 430 | 0.91 | 0.43; 1.34 | 0.788 |
| T2 | 449 | 1.40 | 0.81; 2.41 | 0.231 | 458 | 1.87 | 1.15; 3.10 | **0.013** |
| *NICU* | | | | | | | | |
| T1 | 430 | 0.85 | 0.57; 1.28 | 0.446 | 430 | 0.91 | 0.52; 1.26 | 0.699 |
| T2 | 449 | 0.90 | 0.61; 1.31 | 0.576 | 458 | 1.14 | 0.81; 1.63 | 0.467 |
| *COMPOSITE* | | | | | | | | |
| T1 | 430 | 0.84 | 0.38; 1.86 | 0.670 | 430 | 0.99 | 0.37; 1.43 | 0.987 |
| T2 | 449 | 0.98 | 0.46; 2.07 | 0.968 | 458 | 1.45 | 0.77; 2.78 | 0.262 |

T1, T2, SGA, NICU, COMPOSITE, neonatal adverse outcome (including shoulder dystocia, nerve injury, and bone fracture). Adjusted for maternal age and pre-gestational BMI. $p<0.05$ in boldface.

**Supplementary Table 5.** Plasma GA (KQT), GA (KLV) and GA-total means in early and mid-late pregnancy by GDM status.

| | **GDM** | | **Non-GDM controls** | | |
|---|---|---|---|---|---|
| | *Mean* | *95% CI* | *Mean* | *95% CI* | *P-value*[a] |
| **T1 (N = 436)** | | | | | |
| GA (KQT) (%) | 6.16 | 5.97-6.34 | 6.44 | 6.35-6.52 | 0.563 |
| GA (KLV) (%) | 1.31 | 1.28-1.34 | 1.37 | 1.35-1.38 | 0.067 |
| GA-total (%) | 7.46 | 7.25-7.67 | 7.80 | 7.71-7.90 | 0.423 |
| **T2 (N = 455)** | | | | | |
| GA (KQT) (%) | 5.77 | 5.61-5.93 | 5.96 | 5.89-6.03 | 0.712 |
| GA (KLV) (%) | 1.25 | 1.22-1.28 | 1.26 | 1.24-1.27 | 0.106 |
| GA-total (%) | 7.02 | 6.83-7.20 | 7.21 | 7.13-7.29 | 0.568 |

GA measurements are presented as the mean (95% CI).

[a]Differences of GA means between GDM case and control participants at each visit were assessed using linear mixed-effect regression models adjusted for family history of diabetes, nulliparity, pre-gestational BMI, maternal age at delivery, and gestational age at blood sampling with a participant-specific random intercept.

**Supplementary Fig. 1.**

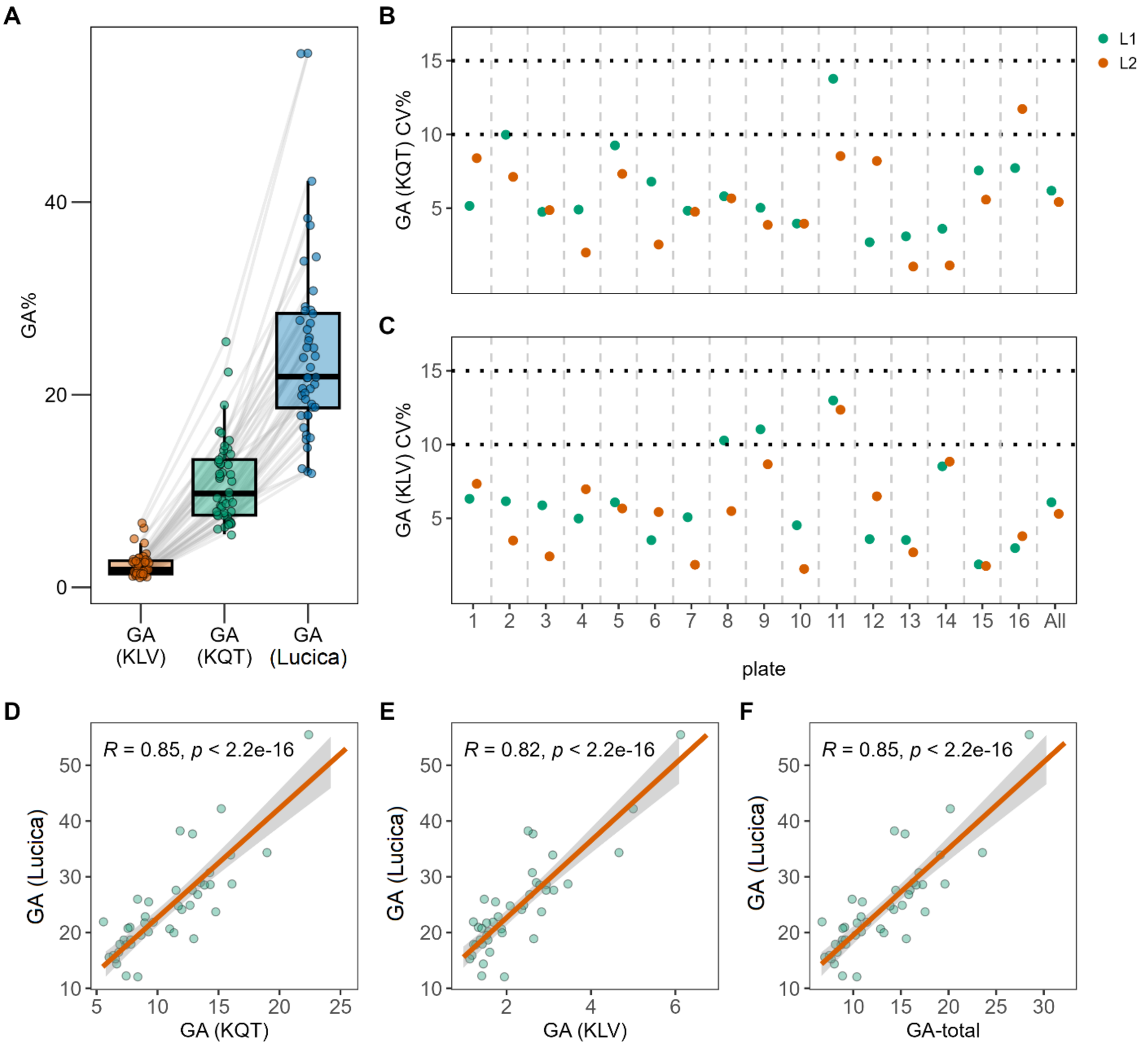


**ESM Fig. 1.** Comparison of GA measurements across methods and plates. A) Distribution of GA (%) measured using the KLV, KQT and Lucica approaches. Individual observations are shown as points, with box plots indicating the median and interquartile range; grey lines connect paired measurements from the same sample across methods. (B, C) CV% for each plate (1-16), for both the KQT (B) and KLV (C) approaches. Green and orange points represent L1 and L2, respectively. Horizontal dotted lines indicate the 10% and 15% reference levels. (D-F) Pairwise relationships between GA measurements obtained with Lucica and GA (KQT) (D), Lucica and GA (KLV) (E), and Lucica and the combined GA-total measure (F). Each point represents an individual sample; the solid line denotes the fitted linear regression and the shaded area the corresponding 95% confidence interval. Spearman correlation coefficients (R) and *p* values are shown within each panel.

**Supplementary Fig. 2.**

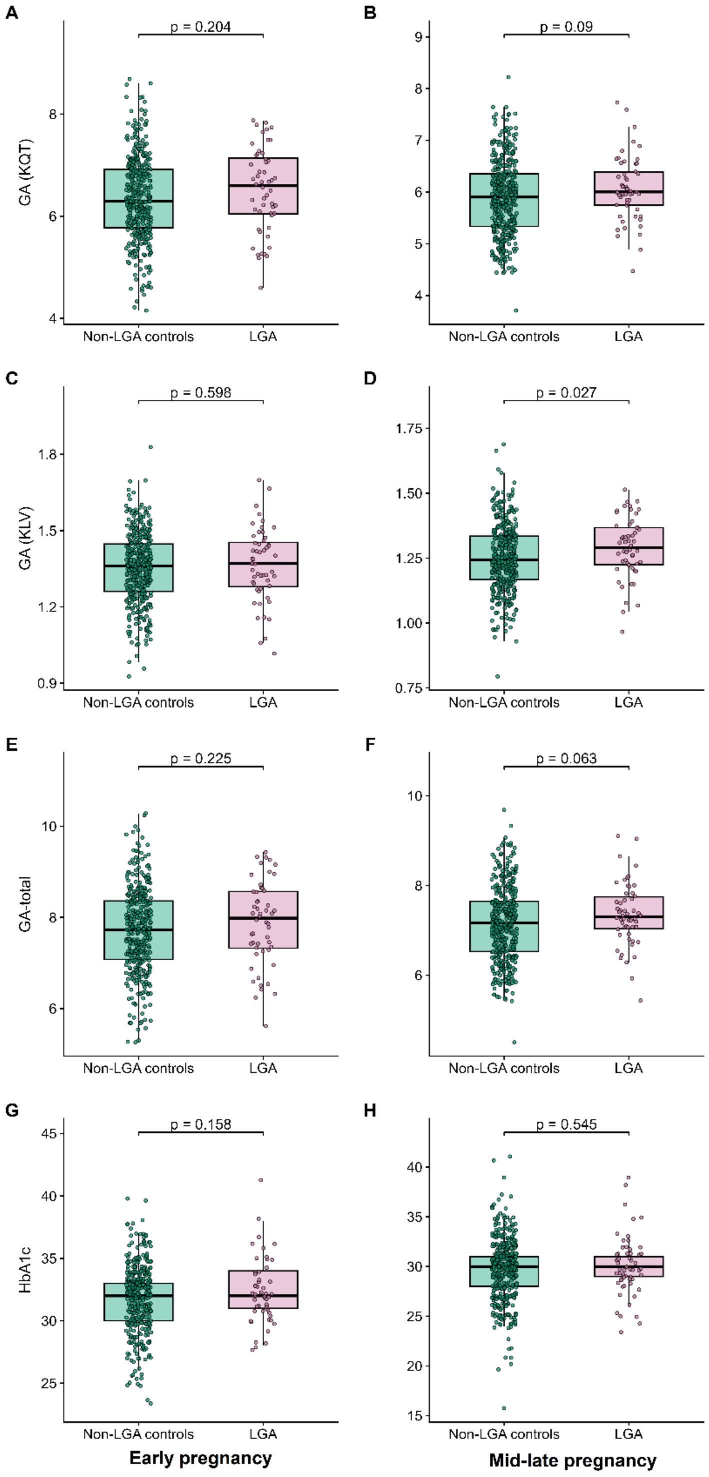


**ESM Fig. 2.** Boxplots showing the distributions of (A, B) GA (KQT), (C, D) GA (KLV), (E, F) GA-total and (G, H) $HbA_{1c}$ in early (left) and mid-late (right) pregnancy between LGA case (pink) and non-LGA controls (green). Between-group differences were assessed using the Wilcoxon rank-sum test for mid-late pregnancy $HbA_{1c}$, owing to its non-normal distribution, and two-sample t-tests for all other measures.
